# EEG frontal alpha asymmetry mediates the association between maternal and child internalizing symptoms in childhood

**DOI:** 10.1101/2024.09.15.24313329

**Authors:** Dashiell D. Sacks, Yiyi Wang, Asja Abron, Kaitlin M. Mulligan, Caroline M. Kelsey, Wanze Xie, Charles A. Nelson, Michelle Bosquet Enlow

**Affiliations:** Department of Psychiatry and Behavioral Sciences, Boston Children’s Hospital, Boston, MA; Department of Psychiatry, Harvard Medical School, Boston, MA; Department of Psychology, University of Chicago, Chicago, IL; Division of Developmental Medicine, Boston Children’s Hospital, Boston, MA; Department of Pediatrics, Harvard Medical School, Boston, MA; School of Psychological and Cognitive Sciences, Peking University, Beijing, China; IDG/McGovern Institute for Brain Research, Peking University, China; Harvard Graduate School of Education, Cambridge, MA

**Keywords:** maternal depression, maternal anxiety, frontal alpha asymmetry, internalizing symptoms, EEG

## Abstract

**Background:** Anxiety and depression are highly prevalent in youth and can cause significant distress and functional impairment. The presence of maternal anxiety and depression are well-established risk factors for child internalizing psychopathology, yet the responsible mechanisms linking the two remain unclear.

**Methods:** We examined the potential mediating and moderating roles of EEG frontal alpha asymmetry (FAA) in the intergenerational transmission of internalizing symptoms in a longitudinal sample of *N* = 323 mother-child dyads. Self-report maternal internalizing symptoms were evaluated at child age 3 years and 5 years, child EEG at 5 years, and parent-report child internalizing symptoms at age 7 years. Mediation was evaluated via bootstrapped (*N* = 5000) confidence intervals.

**Results:** We found significant associations among maternal internalizing (anxiety, depressive) symptoms at child ages 3 and 5 years, child FAA at age 5 years, and child internalizing symptoms at age 7 years. There was a significant mediation effect, whereby greater maternal anxiety and depressive symptoms at age 3 years were significantly associated with greater relative right FAA in children at age 5 years, which, in turn, was significantly associated with greater child internalizing symptoms at age 7 years (*ps*<.001). There was no moderating effect of FAA on the association between maternal internalizing symptoms at age 5 years and child internalizing symptoms at age 7 years.

**Conclusions:** Greater right frontal asymmetry may be a neurophysiological mechanism that mediates the intergenerational transmission of internalizing symptoms.

---

Internalizing disorders (i.e., anxiety and depression) are highly prevalent in youth, with epidemiological studies estimating rates of 11.6% and 12.9%, respectively (Lu, 2019; Tiirikainen et al., 2019). Recent meta-analytic work suggests that these rates approximately doubled to 20.5% and 25.2%, respectively, after the COVID-19 pandemic, emphasizing the need for effective intervention and prevention efforts (Racine et al., 2021). Internalizing disorders, particularly anxiety, have been shown to emerge as early as the preschool years (Bufferd et al., 2011; Egger & Angold, 2006); moreover, early age of onset is associated with long-term chronicity and negative effects on socioemotional functioning (Beesdo et al., 2009; Vergunst et al., 2023).

Maternal internalizing symptoms convey a significant risk for the development of child internalizing problems (Barker et al., 2011; Beidel & Turner, 1997; Goodman et al., 2011). Various studies have explored the impact of timing, with some research indicating that exposure to maternal internalizing symptoms in early childhood (2–3 years) is particularly detrimental (Hentges et al., 2020). The brain is highly vulnerable to environmental stressors during the first years of life, as significant postnatal development and maturation is ongoing (Chen & Baram, 2016). Maternal input is one of the most salient environmental influences during early development (Chen & Baram, 2016). Thus, research is required to elucidate how maternal internalizing symptoms may impact the developing brain and potentially contribute to the development of internalizing symptoms in childhood.

Frontal EEG alpha asymmetry (FAA), which indexes differences in cortical activation in one hemisphere compared to the other at baseline, may index one mechanism involved in the intergenerational transmission of internalizing symptoms. Alpha activity is believed to be inhibitory, with higher alpha activity linked to reduced brain activation. Given this, right FAA refers to right hemispheric power being reduced relative to left power (i.e., a negative FAA value), and left FAA refers to left power being reduced relative to right power (i.e., a positive FAA value) (Vincent et al., 2021). The extant literature indicates that relative FAA is associated with approach versus withdrawal behaviors (Harmon-Jones & Gable, 2018). Specifically, right FAA is associated with greater withdrawal motivation and the processing of negative affect, whereas left FAA is associated with greater approach motivation and the processing of positive affect. Motivational and emotional processing are believed to be lateralized in the brain, with the frontal cortex playing a major role (Harmon-Jones & Gable, 2018). In adults, associations between right FAA and internalizing symptoms are well documented, with research (Henriques & Davidson, 1990) spanning over 30 years.

Studies have also reported associations between right FAA and both anxiety (e.g., Demerdzieva & Pop-Jordanova, 2015) and depression (e.g., Gatzke-Kopp et al., 2014) in children. However, in a meta-analysis by Peltola et al. (2014), which included 20 studies on the association between right FAA and internalizing symptoms in children ranging in age from infancy to adolescence, the effect size was small and not significant overall (*d* = .19, *p* = .08), with eight of the studies reaching significance within the meta-analysis. A key limitation of the individual studies included is the relatively small sample sizes (mean = 64.55; range: 24–135 children). Furthermore, multiple methods for calculating FAA have been used, with different methods linked to different results (Vincent et al., 2021). Results from studies published after this meta-analysis have reported similar mixed evidence for associations (Blaisdell et al., 2020; Feldmann et al., 2018; Grünewald et al., 2018).

Studies have also examined associations between child FAA and maternal internalizing symptoms assessed prenatally (e.g., Diego et al., 2004; Diego et al., 2006; Field et al., 2004) and postnatally (e.g., Forbes et al., 2008; Goldstein et al., 2016; Lopez-Duran et al., 2012). Overall, evidence is suggestive of links between maternal internalizing problems and greater right child FAA. The meta-analysis by Peltola et al. (2014) included 20 studies that investigated associations between psychosocial risk, predominantly maternal depression, and child FAA and reported a significant overall association, with a medium effect size (*d* = .36, *p* < .01). Most studies that have investigated maternal internalizing symptoms and child FAA have focused on maternal depression. More research is needed to understand the effects of maternal anxiety on child FAA. Relatedly, Hill et al. (2020) investigated associations between FAA in 12-month-old infants and in their mothers and observed significant associations, suggesting intergenerational transmission of FAA. Together, these findings suggest that FAA may play a role in the intergenerational transmission of internalizing problems.

The Adaptive Calibration Model (ACM) provides an integrative framework for evaluating how children may adapt to early life stress. Specifically, children’s neural responsivity to stressors is hypothesized to adapt in response to stress exposures, such as maternal anxiety or depression. In the context of FAA, right FAA may reflect neural adaptation to environments characterized by maternal internalizing problems, which may directly increase the risk of child internalizing problems (i.e., a mediating effect). Another possibility is that greater withdrawal motivation, indexed by right FAA, interacts with maternal psychopathology exposure to increase vulnerability and magnify risk (i.e., a moderating effect). Limited indirect evidence supports a potential moderating effect of FAA. For example, Forbes et al. (2008) found that maternal depressive symptoms and child negative affectivity were associated in children with right FAA; child internalizing problems specifically were not measured. Hernandez et al. (2024) investigated potential mediating and moderating effects of FAA in a longitudinal design, which assessed maternal internalizing symptoms (anxiety and depression collapsed into one variable) in pregnancy and child FAA and internalizing symptoms assessed concurrently in 5- to 11-year-old children (*M*age = 7.27). However, they did not find any significant associations of maternal or child symptoms with child FAA. In the extant literature, most studies have been limited by relatively small, cross-sectional samples that do not allow for disentangling of mediating or moderating effects. More research is needed to explicate whether and how FAA may play a role in the association between maternal and child internalizing problems.

The overall goal of the current study was to synthesize these areas of research to examine the potential role of child FAA in the intergenerational transmission of internalizing problems in early to middle childhood. Here, we aimed to investigate associations among maternal internalizing symptoms, child FAA, and child internalizing symptoms in a large longitudinal sample of children. Specifically, we investigated potential mediating and moderating effects of child FAA on the association between maternal and child internalizing symptoms. We considered both maternal anxiety and depressive symptoms, despite common comorbidity, as they may have differential effects on parenting behavior, and, consequently, on child outcomes (Quigley et al., 2023). We hypothesized that 1) greater maternal internalizing symptoms at child ages 3 years and 5 years are associated with greater child relative right FAA at 5 years and greater child internalizing symptoms at age 7 years; 2) child relative right FAA at 5 years mediates the association between maternal internalizing symptoms at 3 years and child internalizing symptoms at 7 years; and 3) child relative right FAA at 5 years moderates the association between maternal internalizing symptoms at 5 years and child internalizing symptoms at 7 years, such that children who demonstrate greater relative right FAA and exposure to heightened maternal internalizing symptoms have the highest level of internalizing symptoms.

## Methods

### Participants

Participants were recruited from a registry of local births comprising families that had indicated willingness to participate in developmental research. Families in the current analyses participated in a prospective longitudinal study to examine the early development of emotion processing. Exclusion criteria included known prenatal or perinatal complications, maternal use of medications during pregnancy that may have a significant impact on fetal brain development (i.e., anticonvulsants, antipsychotics, opioids), pre- or post-term birth (±3 weeks from due date), developmental delay, uncorrected vision difficulties, and neurological disorder or trauma. After enrollment, families were no longer followed, and their data were excluded from analyses if the child was diagnosed with an autism spectrum disorder or a genetic or other condition known to influence neurodevelopment. Families were included in the current analyses if they had maternal internalizing symptom data at child age 3 years and/or 5 years (*n* = 283), child EEG data at 5 years (*n* = 161), and/or child internalizing symptom data at age 7 years (*n* = 213), for an analytic sample of *N* = 323 mother-child dyads.

### Procedures

Mothers were asked to complete questionnaires via an online survey prior to or during laboratory visits. Questionnaires relevant to the current analyses included assessments of sociodemographic characteristics collected at enrollment in infancy, maternal anxiety and depressive symptoms (3 years and 5 years), and child internalizing symptoms (7 years). EEG was collected during the 5-year laboratory visit. The Institutional Review Board at Boston Children’s Hospital approved all methods and procedures used in this study. Parents provided written informed consent prior to the initiation of study activities.

### Measures

#### Demographics

Sociodemographic characteristics were collected via parent report. Data collected included child age at each visit, sex assigned at birth (hereafter “sex”), ethnicity, race, parent education, and annual household income (Table 1).

**Table 1.**
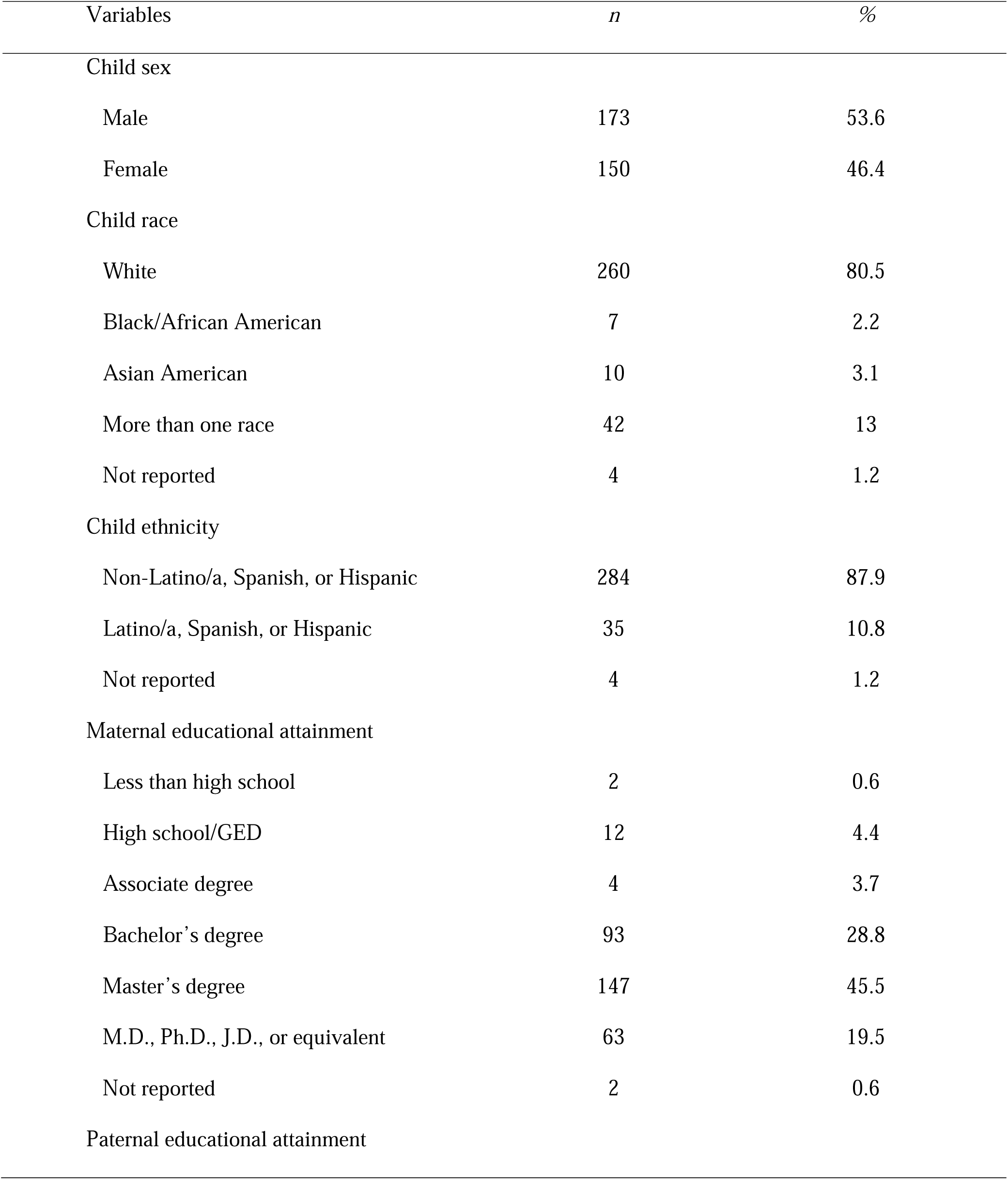
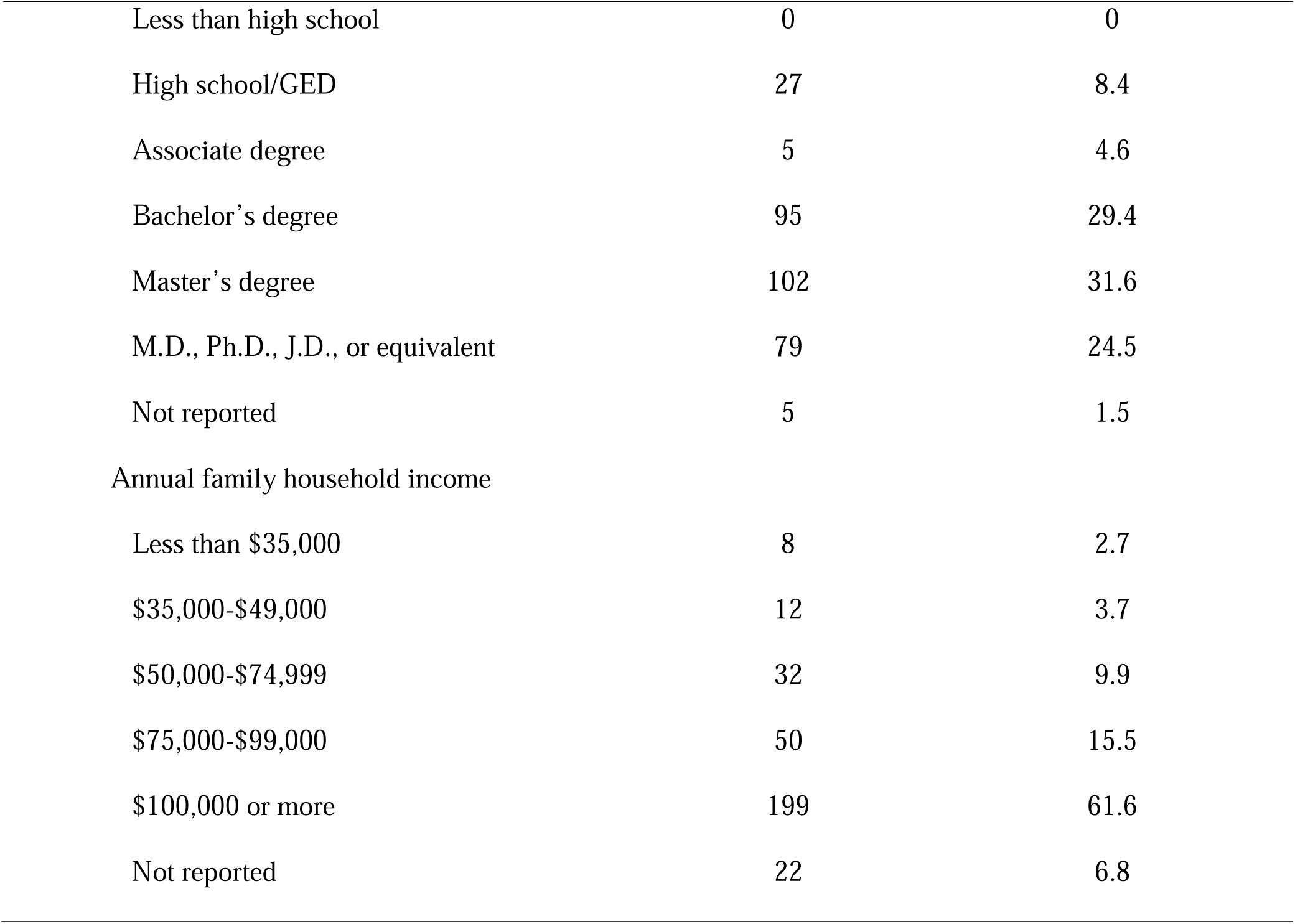
Sample sociodemographic characteristics, collected at infancy (N=323).

| Variables | *n* | % |
| --- | --- | --- |
| Child sex |  |  |
| Male | 173 | 53.6 |
| Female | 150 | 46.4 |
| Child race |  |  |
| White | 260 | 80.5 |
| Black/African American | 7 | 2.2 |
| Asian American | 10 | 3.1 |
| More than one race | 42 | 13 |
| Not reported | 4 | 1.2 |
| Child ethnicity |  |  |
| Non-Latino/a, Spanish, or Hispanic | 284 | 87.9 |
| Latino/a, Spanish, or Hispanic | 35 | 10.8 |
| Not reported | 4 | 1.2 |
| Maternal educational attainment |  |  |
| Less than high school | 2 | 0.6 |
| High school/GED | 12 | 4.4 |
| Associate degree | 4 | 3.7 |
| Bachelor's degree | 93 | 28.8 |
| Master's degree | 147 | 45.5 |
| M.D., Ph.D., J.D., or equivalent | 63 | 19.5 |
| Not reported | 2 | 0.6 |
| Paternal educational attainment |  |  |
| Less than high school | 0 | 0 |
| High school/GED | 27 | 8.4 |
| Associate degree | 5 | 4.6 |
| Bachelor's degree | 95 | 29.4 |
| Master's degree | 102 | 31.6 |
| M.D., Ph.D., J.D., or equivalent | 79 | 24.5 |
| Not reported | 5 | 1.5 |
| Annual family household income |  |  |
| Less than \$35,000 | 8 | 2.7 |
| \$35,000-\$49,000 | 12 | 3.7 |
| \$50,000-\$74,999 | 32 | 9.9 |
| \$75,000-\$99,000 | 50 | 15.5 |
| \$100,000 or more | 199 | 61.6 |
| Not reported | 22 | 6.8 |

#### Maternal Anxiety Symptoms (3 years, 5 years; Predictor)

Maternal anxiety symptoms were measured at child ages 3 years and 5 years via the Trait Anxiety form of the Spielberger State-Trait Anxiety Inventory (STAI-T; (Spielberger et al., 1983)). The STAI is a 20-item self-report questionnaire designed to measure anxiety proneness. Respondents are asked to rate the frequency of general mood states on a 4-point scale, ranging from ‘almost never’ to ‘almost always.’ Item scores were summed to create a total score (possible range: 20 – 80), with a higher score indicating greater anxiety. The STAI-T has established good internal consistency (α = .90), and test-retest coefficients from .73 to .86 (Barnes et al., 2002). Cronbach’s alpha scores in this sample were .89 and .91 at ages 3 years and 5 years, respectively.

#### Maternal Depressive Symptoms (3 years, 5 years; Predictor)

Maternal depressive symptoms were measured at child ages 3 years and 5 years via the Beck Depression Inventory (BDI-IA; (Beck et al., 1961)). The BDI is a 21-item self-report questionnaire that assesses the frequency and intensity of depressive symptoms in the past week.

Items are scored on a 4-point scale (range 0-3) and summed to create a total score, with a total possible range of 0 – 63; higher scores indicate greater depressive symptoms. Internal consistency for the BDI has been reported as ranging from .73 to .92, with a mean of .86 (Beck et al., 1988). Cronbach’s alpha scores in this sample were .77 and .84 at ages 3 years and 5 years, respectively.

#### Child Internalizing Symptoms (7 years; Outcome)

Mothers completed the Child Behavior Checklist 6-18 (CBCL/6-18) at the 7-year assessment. The CBCL forms are well established, empirically supported questionnaires for assessing child psychopathology symptoms (Achenbach & Rescorla, 2000; Achenbach et al., 2001; Achenbach & Edelbrock, 1991; Achenbach & Rescorla, 2014). The 113-item CBCL/6-18 produces scores on multiple syndrome and DSM-oriented scales, as well as higher-order symptom scores. Parents are asked to report on their children’s behavior during the past six months, with possible item scores ranging from 0 (‘not true’) to 2 (‘very true or often true’). The current analyses focused on the Internalizing Problems scale, which comprises the following syndrome scales: Anxious/Depressed, Somatic Complaints, and Withdrawn. Subscale raw scores are calibrated and normed by child age and gender, with normed scores expressed as the standard T-score metric. T-scores < 60 suggest non-clinical level of symptoms, 60-63 borderline clinical significance, and ≥ 64 clinical significance. Cronbach’s alpha in this sample was .89.

#### Child Frontal Alpha Asymmetry (5 years; Mediator/Moderator)

##### EEG Acquisition

Continuous scalp EEG was recorded at 5 years using a 128-electrode HydroCel Geodesic Sensor Net (HGSN; Electrical Geodesic Inc.). The net was connected to a NetAmps 300 amplifier (Electrical Geodesic Inc.) and referenced online to a single vertex electrode (Cz). The placement of the net was checked and adjusted by the experimenter, and channel impedances were kept at or below 50 kΩ, Baseline EEG was collected and recorded with a sampling rate of 500 Hz for 4 minutes while children watched a silent, nonsocial, ‘screensaver style’ video showing patterned lights.

##### Preprocessing

The EEG data were preprocessed using the EEGLAB (Delorme & Makeig, 2004) and ERPLAB (Lopez-Calderon & Luck, 2014) toolboxes in MATLAB (R2022a, the Mathworks, Inc.). Continuous EEG data were filtered with an eighth order Butterworth bandpass filter with a pass band of 1−50 Hz and segmented into 2s epochs. Bad channels were detected and removed using the FASTER EEGLAB plugin (i.e., an absolute Z score greater than 3). Independent component analysis (ICA) was performed to remove components related to eye movements, blinks, and focal activity using the ADJUST script (Mognon et al., 2011). The EEG epochs were further evaluated for artifacts using absolute threshold (EEG > 100 µV or EEG < −100 µV) and frequency threshold (performed on 20 to 50HZ with a threshold between −100 to 30 dB). Channel interpolation was conducted using a spherical spline interpolation if there were fewer than 18 (15%) electrodes that were missing or had bad data; otherwise, the epoch was excluded. The mean number of epochs retained among the sample was 113.86, *SD* = 9.15.

##### Power Analysis and FAA Calculation

Power spectral density (PSD) was calculated using the Fieldtrip toolbox (Oostenveld et al., 2011). Fast Fourier transform was applied to EEG epochs with a 0.5s width Hanning window. The PSD was calculated for all frequency bins from 1 to 30 Hz. The boundary of the alpha band was defined based on individual alpha peak frequency (IAF; (*M* = 8.60, *SD* = 0.59, range 7.5 to 10.5). The alpha band for each individual was generated using the proportion of the IAF, .8*IAF, 1.2*IAF (Perone et al., 2018).

FAA was calculated using the method recommended for longitudinal research by Vincent et al. (2021), as it showed the most stability over time. Relative alpha power was calculated as the proportion of alpha PSD relative to the total PSD from the beginning of the child theta band to 30 Hz (i.e., 3−30 Hz). The log relative alpha power was averaged over the electrodes including and surrounding the F3 (19, 20, 23, 24, 27, and 28) and F4 (3, 4, 117, 118, 123, and 124) 10−20 positions (Figure 1). A negative asymmetry value indicates right frontal asymmetry (withdrawal motivation), and a positive asymmetry value indicates left frontal asymmetry (approach motivation).

**Figure 1.**
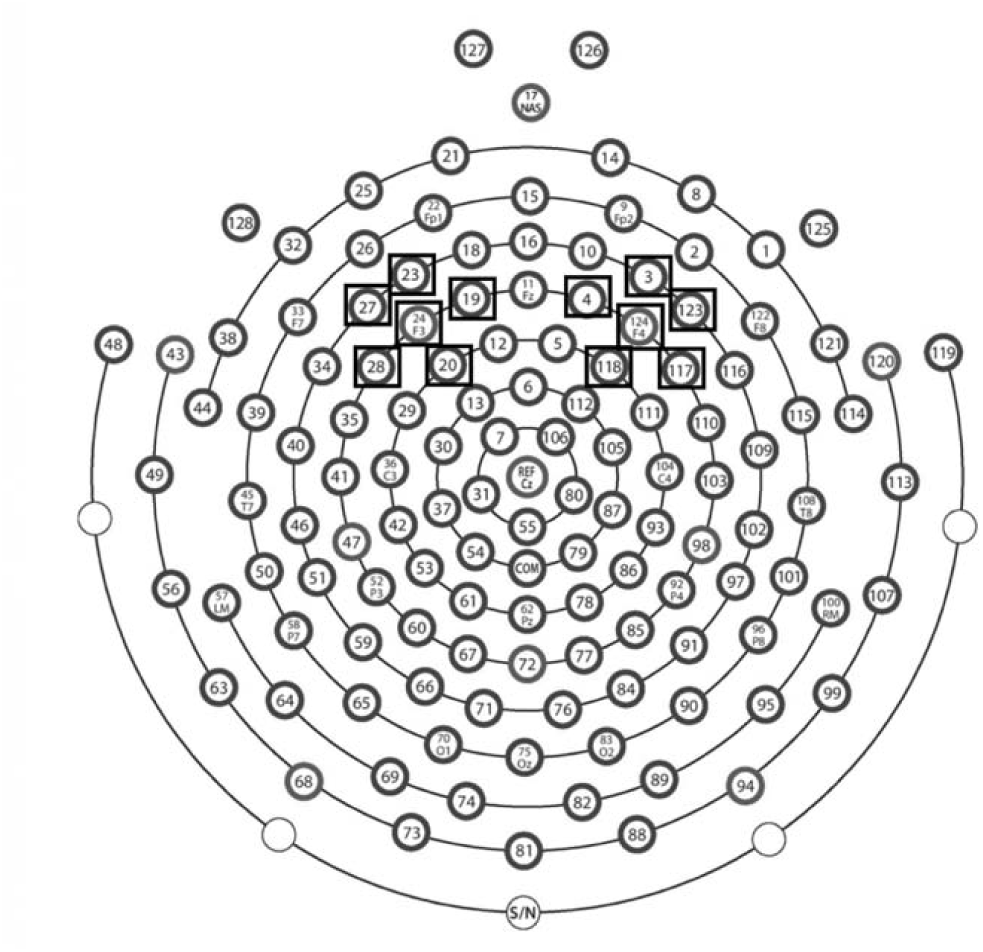
Schematic of a 128-channel HydroCel Geodesic Sensor Net. Electrodes used for the left and right regions of interest (ROIs) to assess frontal alpha asymmetry (FAA) are in black boxes.

### Statistical Analysis

Statistical analyses were conducted in Python 3. Descriptive statistics for the sample sociodemographic characteristics and main study variables were calculated. Pearson’s correlations were conducted to examine bivariate associations among the main study variables and sociodemographic characteristics. Mediation and moderation analyses were conducted using pyprocessmacro (https://github.com/QuentinAndre/pyprocessmacro), a Python implementation of the PROCESS Macro (André, 2021). Separate mediation models were investigated with maternal anxiety or depressive symptoms at child age 3 years as the predictor, FAA at child age 5 years as the mediator, and child internalizing symptoms at child age 7 years as the outcome. Individual paths and direct, indirect, and total effects were examined for each model. Significance of indirect effects was determined using bootstrapped (*n* = 5000) confidence intervals (95% CI). Additionally, two moderation models examined the main effects of maternal anxiety or depressive symptoms at child age 5 years and FAA at child age 5 years and the interaction effect of maternal symptoms and child FAA on child internalizing symptoms at 7 years. For the mediation and moderation analyses, missing data at each timepoint were imputed using MissForest (https://github.com/yuenshingyan/MissForest). For each of these analyses, a supplementary analysis was conducted for model comparison in the subset of participants with complete data for all the model variables (*N* = 97 for mediation, *N* = 111 for moderation).

## Results

### Sample Characteristics and Descriptive Analyses

Sample sociodemographic characteristics are presented in Table 1. Children were predominantly non-Hispanic White, and parental education and annual household income indicated middle to high socioeconomic status. Descriptive statistics for the main study variables are presented in Table 2. These variables were not associated with any of the sociodemographic variables; thus, these variables were not considered further in analyses. Bivariate associations among the main study variables are presented in Table 3. There were significant associations between all main study variables except for maternal depressive symptoms at 5 years and child FAA at 5 years (*p* = .11).

**Table 2.** Descriptive statistics for the main study variables.

|  | Min | Max | Mean | SD |
| --- | --- | --- | --- | --- |
| Maternal anxiety symptoms, 3 years | 21 | 57 | 33.23 | 6.94 |
| Maternal anxiety symptoms, 5 years | 20 | 58 | 33.00 | 7.90 |
| Maternal depressive symptoms, 3 years | 0 | 27 | 5.15 | 4.11 |
| Maternal depressive symptoms, 5 years | 0 | 24 | 4.97 | 4.52 |
| Frontal alpha asymmetry (FAA), 5 years | -.291 | .360 | .002 | .131 |
| Child internalizing T-score, 7 years | 33 | 72 | 47.10 | 9.73 |

*Note.* Maternal anxiety symptoms calculated from the Trait anxiety form of the Spielberger State-Trait Anxiety Inventory (STAI-T); maternal depressive symptoms calculated from the Revised Beck Depression Inventory (BDI-IA); child internalizing T-score from the Child Behavior Checklist 6-18 years (CBCL) Internalizing Problems Scale.

**Table 3.** Bivariate associations among the main study variables.

|  | STAI – 3Y | STAI – 5Y | BDI –3Y | BDI –5Y | FAA – 5Y |
| --- | --- | --- | --- | --- | --- |
| STAI – 5 years | **.74<sup>\*\*\*</sup>** | -- |  |  |  |
| BDI –3 years | **.58<sup>\*\*\*</sup>** | **.39<sup>\*\*\*</sup>** | -- |  |  |
| BDI –5 years | **.48<sup>\*\*\*</sup>** | **.65<sup>\*\*\*</sup>** | **.59<sup>\*\*\*</sup>** | -- |  |
| FAA – 5 years | **-.23<sup>\*\*</sup>** | **-.16<sup>\*</sup>** | **-.18<sup>\*</sup>** | -.13 | -- |
| CBCL – 7 years | **.31<sup>\*\*\*</sup>** | **.27<sup>\*\*\*</sup>** | **.32<sup>\*\*\*</sup>** | **.27<sup>\*\*\*</sup>** | **-.29<sup>\*\*</sup>** |

*Note.* STAI = Trait Anxiety form of the Spielberger State-Trait Anxiety Inventory (maternal anxiety symptoms); BDI = Revised Beck Depression Inventory (maternal depressive symptoms); FAA = EEG frontal alpha asymmetry (log relative); CBCL = Internalizing Problems T-score from the Child Behavior Checklist 6-18 years (child internalizing symptoms).
\* $p < .05$ , \*\* $p < .01$ , \*\*\* $p < .001$

### Mediation Analyses

#### Maternal Anxiety Symptoms

A mediation analysis was conducted to examine whether child FAA at age 5 years mediated the association between maternal anxiety symptoms at child age 3 years and child internalizing symptoms at age 7 years. Each pathway was significant (Table 4). Specifically, greater maternal anxiety was significantly associated with reduced FAA (*p*< .001), which was significantly associated with greater child internalizing symptoms (*p*< .001). The indirect effect of maternal anxiety on child internalizing symptoms through FAA was significant 95% CI [0.052, 0.166]. The direct effect of maternal anxiety on child internalizing symptoms remained significant when controlling for FAA (*p*< .001). These findings indicate that FAA at age 5 years mediated the association between maternal anxiety symptoms at age 3 years and child internalizing symptoms at age 7 years (Figure 2).

**Figure 2.**
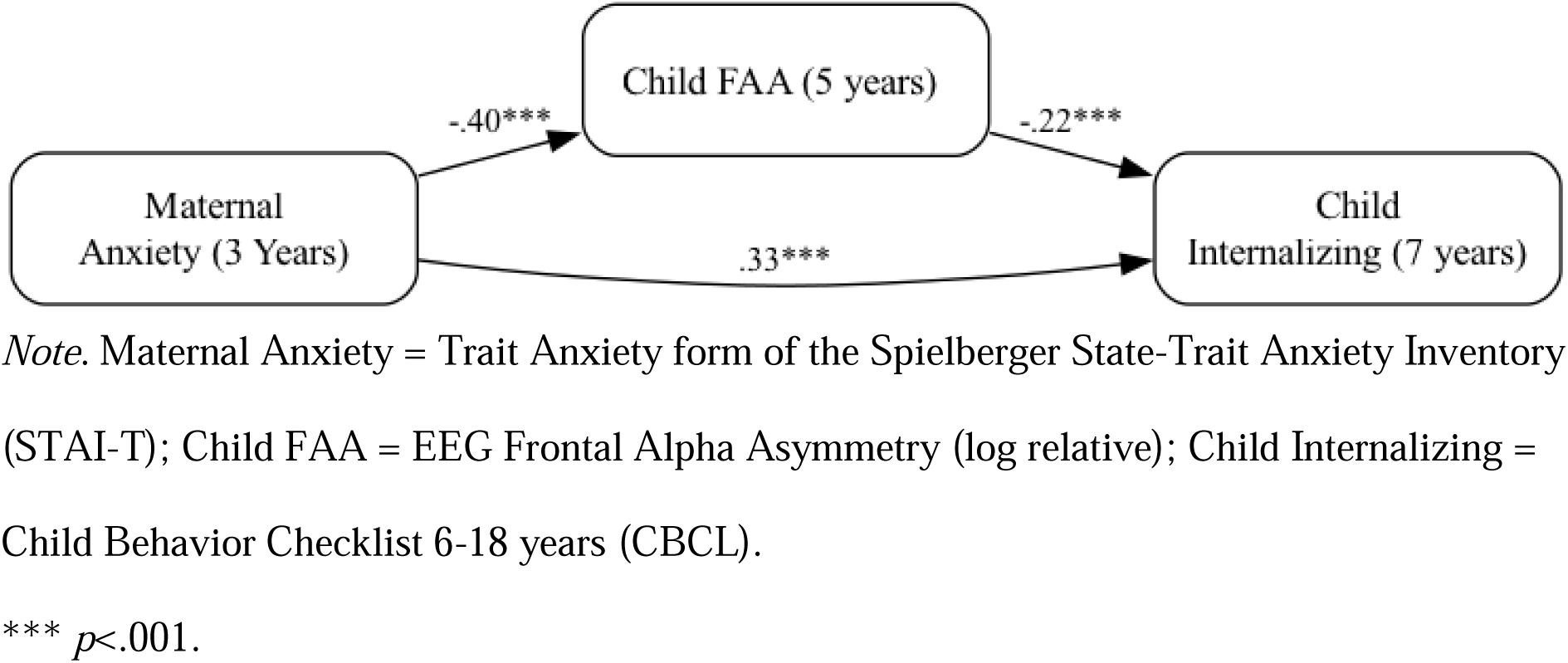
Mediation model for the association between maternal anxiety symptoms at 3 years and child internalizing symptoms at 7 years, mediated by child FAA at 5 years.

**Table 4.** Mediation models for the association between maternal anxiety/depressive symptoms at 3 years and child internalizing symptoms at 7 years, mediated by child FAA at 5 years.

| Pathway | Measures | B | B(S) | SE | t | Sig. | 95% CI |  |
| --- | --- | --- | --- | --- | --- | --- | --- | --- |
|  |  |  |  |  |  |  | LLCI | ULCI |
| Maternal Anxiety |  |  |  |  |  |  |  |  |
| IV → M | STAI → FAA | -0.01 | -.40 | 0.001 | -7.62 | < .001 | -0.008 | -0.005 |
| M → DV | FAA → CBCL | -17.66 | -.22 | 4.27 | -4.13 | < .001 | -26.03 | -9.285 |
| IV → DV | STAI → CBCL | 0.40 | .33 | 0.06 | 6.15 | < .001 | 0.271 | 0.524 |
| (Direct Effect) |  |  |  |  |  |  |  |  |
| IV → M → DV | STAI → FAA | 0.11 | .09 | .03 |  |  | 0.052 | 0.166 |
| (Indirect Effect) | → CBCL |  |  |  |  |  |  |  |
| Maternal Depression |  |  |  |  |  |  |  |  |
| IV → M | BDI → FAA | -0.01 | -.33 | 0.001 | -6.30 | < .001 | -0.011 | -0.006 |
| M → DV | FAA → CBCL | -19.12 | -.24 | 4.13 | -4.63 | < .001 | -27.224 | -11.025 |
| IV → DV | BDI → CBCL | 0.68 | .34 | 0.11 | 6.51 | < .001 | 0.477 | 0.888 |
| (Direct Effect) |  |  |  |  |  |  |  |  |
| IV → M → DV | BDI → FAA | 0.16 | .08 | 0.05 |  |  | 0.077 | 0.271 |
| (Indirect Effect) | → CBCL |  |  |  |  |  |  |  |

*Note.* STAI = Trait Anxiety form of the Spielberger State-Trait Anxiety Inventory (maternal anxiety symptoms); BDI = Revised Beck Depression Inventory (maternal depressive symptoms); FAA = EEG Frontal Alpha Asymmetry (log relative); CBCL = Internalizing Problems T-score from the Child Behavior Checklist 6-18 years (child internalizing symptoms).

**Table 5.** Regression models for maternal symptoms (5 years), child FAA (5 years), and child internalizing symptoms (7 years)

| Variable | B | B(S) | SE | t | Sig. | 95% CI |  |
| --- | --- | --- | --- | --- | --- | --- | --- |
|  |  |  |  |  |  | LLCI | ULCI |
| Maternal Anxiety |  |  |  |  |  |  |  |
| Maternal Anxiety – 5 Years | 0.33 | .28 | 0.06 | 5.12 | <.001 | 0.20 | 0.45 |
| Child FAA – 5 years | -20.46 | -.26 | 4.31 | -4.75 | <.001 | -28.91 | -12.01 |
| Maternal Anxiety*Child FAA | 0.51 | .05 | 0.55 | 0.92 | .360 | -0.58 | 1.60 |
| Maternal Depression |  |  |  |  |  |  |  |
| Maternal Depression – 5 Years | 0.67 | .35 | 0.10 | 6.54 | <.001 | 0.47 | 0.87 |
| Child FAA – 5 years | -18.73 | -.24 | 4.16 | -4.50 | <.001 | -26.88 | -10.57 |
| Maternal Depression*Child FAA | 01.38 | .08 | 0.87 | 1.57 | .116 | -0.34 | 3.09 |

*Note.* Maternal Anxiety = Trait Anxiety form of the Spielberger State-Trait Anxiety Inventory (STAI-T); Maternal Depression = Revised Beck Depression Inventory (BDI-IA); FAA = EEG Frontal Alpha Asymmetry (log relative); Child Internalizing = Internalizing Problems T-score from the Child Behavior Checklist 6-18 years (CBCL).

#### Maternal Depressive Symptoms

A mediation analysis was conducted to examine whether child FAA at age 5 years mediated the association between maternal depressive symptoms at child age 3 years and child internalizing symptoms at age 7 years. Each pathway was significant (Table 4). Greater maternal depressive symptoms were significantly associated with reduced child FAA (*p*< .001), which was significantly associated with greater child internalizing symptoms (*p*< .001). The indirect effect of maternal depressive symptoms on child internalizing symptoms through FAA was significant 95% CI [0.077, 0.271]. The direct effect of maternal depressive symptoms on child internalizing symptoms remained significant when controlling for FAA (*p*< .001). These findings indicate that FAA at age 5 years mediates the association between maternal depressive symptoms at age 3 years and child internalizing symptoms at age 7 years (Figure 3).

**Figure 3.**
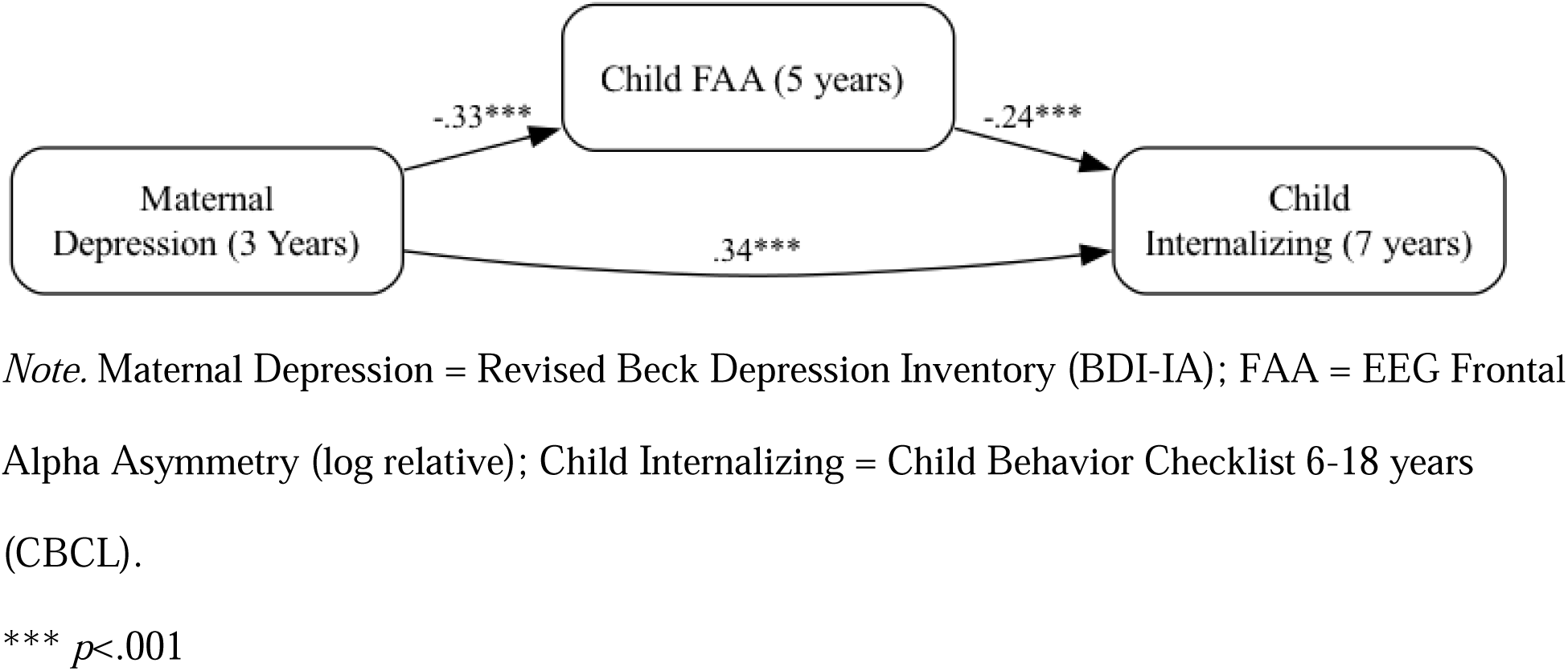
Mediation model for the association between maternal depressive symptoms at 3 years and child internalizing symptoms at 7 years, mediated by child FAA at 5 years.

### Moderation Analysis

Moderation analyses tested whether child FAA at age 5 years moderated the association between maternal internalizing symptoms at 5 years and child internalizing symptoms at 7 years. Across the anxiety and depression models, there were no significant moderation effects. Rather, maternal symptoms and child FAA were independently associated with child symptoms, as detailed below.

#### Maternal Anxiety Symptoms

Child FAA at 5 years and maternal anxiety symptoms at 5 years independently contributed to the prediction of child internalizing symptoms at 7 years (*p*s<.001). The interaction term (child FAA x maternal anxiety symptoms) was not significant (*p* =.360). Thus, no moderation effect was observed.

#### Maternal Depressive Symptoms

Child FAA at 5 years and maternal depressive symptoms at 5 years independently contributed to the prediction of child internalizing symptoms at 7 years (*p*s<.001). The interaction term (child FAA x maternal depressive symptoms) was not significant (*p* =.116). Thus, no moderation effect was observed.

### Supplementary Analyses

In supplementary analyses, the results for all mediation and moderation models remained consistent in the subset of participants with complete data across time points. See Supplementary Materials for full model details.

## Discussion

In this study we examined whether child FAA at age 5 years mediates and/or moderates associations between maternal anxiety or depressive symptoms in early childhood (3 years, 5 years) and child internalizing symptoms at 7 years. We selected these ages given evidence that maternal internalizing symptoms may convey particular risk in early childhood for the development of child internalizing symptoms (Hentges et al., 2020). We observed significant associations among child FAA and maternal and child internalizing symptoms. Moreover, there was a significant mediating effect of right child FAA at 5 years on the association between greater maternal internalizing symptoms at 3 years and greater child internalizing symptoms at 7 years. We did not observe a moderating effect of child FAA at 5 years on the association between maternal internalizing symptoms at 5 years and child internalizing symptoms at 7 years. Rather, there was an additive effect, whereby greater right child FAA at 5 years and maternal internalizing symptoms at 5 years independently contributed to the prediction of greater child internalizing symptoms at 7 years.

To our knowledge, this is the first study to demonstrate that FAA mediates associations between maternal and child internalizing symptoms. More specifically, greater maternal anxiety and depressive symptoms at child age 3 years were each associated with greater right FAA at age 5 years, which, in turn, was associated with greater child internalizing symptoms at age 7 years. These findings suggest that child right frontal alpha asymmetry may be one mechanism through which maternal anxiety and depression increase child vulnerability to internalizing problems. These mediation effects underscore the importance of early neural markers in understanding the transmission of internalizing symptoms from mother to child.

Interest in frontal asymmetry stems from early evidence that motivational and emotional processing is lateralized in the frontal cortex (Davidson & Fox, 1982; Harmon-Jones & Gable, 2018). Exposure to maternal internalizing symptoms may lead to greater development/activation in the right frontal cortex and/or suppression in the left frontal cortex and thus greater overall dominance in rightward asymmetrical activity. These changes may then increase withdrawal motivation and processing of negative affect, which in turn increases risk for internalizing symptoms. In the current analyses, the direct effect of maternal symptoms on child symptoms remained significant even after accounting for FAA, suggesting that neural changes indexed by FAA are only one pathway by which maternal symptoms may influence child symptoms. Continued research in this area is important to delineate further mechanisms by which maternal internalizing psychopathology is transmitted intergenerationally.

We did not find significant moderating effects of FAA on the association between maternal internalizing symptoms at 5 years and child internalizing symptoms at 7 years. Rather, there was an additive effect, whereby both maternal symptoms and child FAA were independently associated with child symptoms. Although some previous work suggests moderating effects of FAA (e.g., Forbes et al. (2008), these studies did not attempt to disentangle mediating and moderating effects longitudinally. Thus, they may not have fully captured the nuanced associations among these variables. Differences in the pattern of findings across studies may also relate to the timing of measurement of each of the constructs in relation to each other (i.e., amount of time between assessments) and within the child’s development (e.g., early childhood vs. later childhood). Future longitudinal research with repeated measures across development will be required to determine any potential timing effects.

To date, most research in this area has focused on maternal depressive psychopathology. Here, we built on existing literature by investigating mediation and moderation models in relation to both maternal anxiety and depressive symptoms. Our findings suggest potential impact of both maternal anxiety and depressive symptoms on offspring FAA and subsequent internalizing symptoms. Future research may consider testing these models in samples with less overlap in symptoms, if disentangling specific effects or relative contributions of maternal anxiety versus depression is of interest. Future research may also consider investigating potential contributory mechanisms (e.g., caregiving behaviors) by which maternal internalizing conditions may impact the developing brain of the child. Finally, these models should be tested in samples with greater clinical severity to determine if the observed associations vary by maternal symptom severity. The current findings suggest that even mild levels of maternal symptoms may impact child FAA and risk for internalizing problems.

Hernandez et al. (2024) did not find a significant association between maternal symptoms and child FAA in their large, longitudinal sample. However, whereas Hernandez et al. (2024) investigated associations between prenatal maternal symptoms and child FAA at a mean age of 7.27 years, we focused on maternal symptoms at 3 years and child FAA at 5 years, a 2-year interval. One potential reason for the differences across studies may be exposure timing effects: The preschool period may be a particularly sensitive period for effects of maternal symptoms on child brain development; moreover, effects may have greater significance more proximally. Furthermore, the intervening mechanisms likely differ between prenatal and postnatal exposure. Whereas prenatal transmission would indicate genetic predisposition (or vulnerability) and/or an adverse prenatal environment, postnatal transmission may also relate to environmental influences, such as caregiving behaviors (Goodman & Gotlib, 1999). Although some studies have reported associations between prenatal symptoms and child FAA (e.g., Diego et al., 2004; Field et al., 2004), they have primarily reported such associations at infancy. Studies that have reported associations at later ages have primarily involved parents with clinical symptomology.

Hernandez et al. (2024) also did not observe a significant association between child FAA and child internalizing symptoms. The extant literature is mixed, with the effect not reaching significance in the meta-analysis by Peltola et al. (2014) (*d* = .19, *p* = .08). Some variation in findings across studies may relate to differences in the methods used for calculating FAA. Here we used the method found to show the greatest longitudinal stability (Vincent et al., 2021). Further, whereas Hernandez et al. (2024) investigated concurrent associations between FAA and symptoms in children aged 5-11 (*M* = 7.27), we investigated predictive associations over a smaller age range. Future longitudinal work should more closely consider timing and age effects in these associations.

### Strengths and Limitations

Strengths of this study include the large, longitudinal sample and repeated measures at three fixed intervals, which allowed us to establish a clear temporal sequence for mediation. Nevertheless, the findings should be considered in the context of the study limitations. The sample comprised families who are predominantly White and of middle to high socioeconomic status, which may reduce the generalizability of the findings. Levels of maternal psychopathology were relatively low; the nature of associations among the main variables could vary in samples with more severe symptoms. Despite these relatively low levels of symptoms, we observed significant mediation effects, suggesting that maternal symptoms may influence child neural development and consequent internalizing behaviors even when present at more mild levels. Finally, the measures of maternal and child internalizing symptoms relied on maternal report, which could introduce bias, leading to inflated correlations. However, the results were mediated by EEG FAA, an objectively recorded biological measure, which likely reduces the impact of such bias on our findings. Moreover, a recent report confirms earlier findings that maternal psychopathology minimally biases maternal reports of child emotional and behavioral problems (Olino et al., 2021).

## Conclusion

This study provides novel insights into the role of FAA in the intergenerational transmission of internalizing symptoms. Specifically, there was a significant mediating effect, whereby maternal anxious and depressive symptoms at child age 3 years were associated with child right FAA at age 5 years, which, in turn, was associated with greater child internalizing symptoms at age 7 years. There was no moderating effect at 5 years. Rather, maternal symptoms and child FAA were independently associated with child symptoms at age 7 years. Overall, the results support a mediating role of FAA, which was suggested in prior studies that investigated bivariate associations, but not previously empirically tested in longitudinal studies. Our findings suggest that FAA may serve as a neurophysiological mechanism in the intergenerational transmission of internalizing behaviors. Future research should continue to investigate the complex interplay between neural markers and environmental influences to enhance our understanding of socioemotional development across childhood. Furthermore, future studies should continue to explore the longitudinal impact of maternal internalizing symptoms on child brain development and mental health, including disentangling potential timing effects of exposure to maternal psychopathology. Our findings support the relevance of intervention programs targeting maternal mental health, particularly in early childhood, given the apparent influence of maternal internalizing symptoms on child brain development during this vulnerable period and subsequent internalizing psychopathology.

## Data Availability

The data that support the findings of this study are available from the corresponding author upon reasonable request.

## Acknowledgements

This research was supported by grants from the National Institute of Mental Health (MH078829) to CAN and MBE and from the Tommy Fuss Center for Neuropsychiatric Disease Research at Boston Children’s Hospital to MBE. Study data were collected and managed using Research Electronic Data Capture (REDCap) tools hosted at Boston Children’s Hospital. We are extremely grateful for the parents and infants who participated in this study, without whom this research would not be possible.

