## Supplement for "EEG frontal alpha asymmetry mediates the association between maternal and child internalizing symptoms in childhood"

**Supplementary Mediation**

### Maternal Anxiety Symptoms

A mediation analysis was conducted to examine whether child FAA at age 5 years mediated the association between maternal anxiety symptoms at child age 3 years and child internalizing symptoms at age 7 years. Each pathway was significant (Supplementary Table 1). Specifically, greater maternal anxiety was significantly associated with reduced FAA (*p* = .002), which was significantly associated with greater child internalizing symptoms (*p*< .041). The indirect effect of maternal anxiety on child internalizing symptoms through FAA was significant 95% CI [0.073, 0.581]. The direct effect of maternal anxiety on child internalizing symptoms remained significant when controlling for FAA (*p*< .013). These findings indicate that FAA at age 5 years mediated the association between maternal anxiety symptoms at age 3 years and child internalizing symptoms at age 7 years (Supplementary Figure 1).

**Supplementary Table 1**

*Mediation model showing that child FAA at age 5 years mediated the association between maternal anxiety symptoms at age 3 years and child internalizing symptoms at age 7 years in subsample with complete data.*

| Pathway | Measures | B | B(S) | SE | t | p | LLCI | ULCI |
| --- | --- | --- | --- | --- | --- | --- | --- | --- |
| IV → M | STAI → FAA | -0.01 | -.34 | 0.002 | -3.14 | .002 | -0.010 | -0.002 |
| M → DV | FAA → CBCL | -12.96 | -.18 | 6.24 | -2.08 | .041 | -25.198 | -0.721 |
| IV → DV (Direct Effect) | STAI → CBCL | 0.33 | .23 | 0.13 | -2.52 | .013 | 0.073 | 0.581 |
| IV → M → DV  (Indirect Effect): | STAI → FAA → CBCL | 0.08 | .06 | 0.05 |  |  | 0.073 | 0.581 |

*Note*. STAI = Trait Anxiety form of the Spielberger State-Trait Anxiety Inventory; FAA = EEG Frontal Alpha Asymmetry (log relative); CBCL = Child Behavior Checklist 6-18 years.

**Supplementary Figure 1**

*Mediation model for the association between maternal anxiety symptoms at 3 years and child internalizing symptoms at 7 years, mediated by child FAA at 5 years in subsample with complete data.*

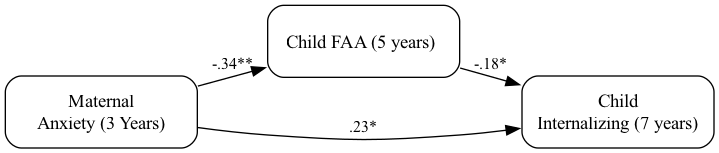

*Note*. Maternal Anxiety = Trait Anxiety form of the Spielberger State-Trait Anxiety Inventory (STAI-T); FAA = EEG Frontal Alpha Asymmetry (log relative); Child Internalizing = Child Behavior Checklist 6-18 years (CBCL).

* *p*<.05, **** *p* <.01.

### Maternal Depressive Symptoms

A mediation analysis was conducted to examine whether child FAA at age 5 years mediated the association between maternal depressive symptoms at child age 3 years and child internalizing symptoms at age 7 years. Each pathway was significant (Supplementary Table 2). Greater maternal depressive symptoms were significantly associated with reduced child FAA (*p* = .0499), which was significantly associated with greater child internalizing symptoms (*p*< .018). The indirect effect of maternal depressive symptoms on child internalizing symptoms through FAA was significant 95% CI [0.006, 0.312]. The direct effect of maternal depressive symptoms on child internalizing symptoms remained significant when controlling for FAA (*p*< .009). These findings indicate that FAA at age 5 years mediates the association between maternal depressive symptoms at age 3 years and child internalizing symptoms at age 7 years (Supplementary Figure 2).

**Supplementary Table 2**

*Mediation model with FAA mediating the association between maternal depressive symptoms at child age 3 years and child internalizing symptoms at 7 years in subsample with complete data.*

| Pathway | Measures | B | B(S) | SE | t | p | LLCI | ULCI |
| --- | --- | --- | --- | --- | --- | --- | --- | --- |
| IV → M | BDI → FAA | -0.01 | -.20 | 0.003 | -1.99 | .0499 | -0.013 | -0.0001 |
| M → DV | FAA → CBCL | -14.58 | -.20 | 6.05 | -2.41 | .018 | -26.425 | -2.729 |
| IV → DV (Direct Effect) | BDI → CBCL | 0.513 | .22 | 0.19 | 2.65 | .009 | 0.134 | 0.891 |
| IV → M → DV  (Indirect Effect): | BDI → FAA → CBCL | 0.093 | .04 | 0.08 |  |  | 0.006 | 0.312 |

*Note.* BDI = Revised Beck Depression Inventory; FAA = EEG Frontal Alpha Asymmetry (log relative); CBCL = Child Behavior Checklist 6-18 years.

**Supplementary Figure 2**

*Mediation model for the association between maternal depressive symptoms at 3 years and child internalizing symptoms at 7 years, mediated by child FAA at 5 years in subsample with complete data.*

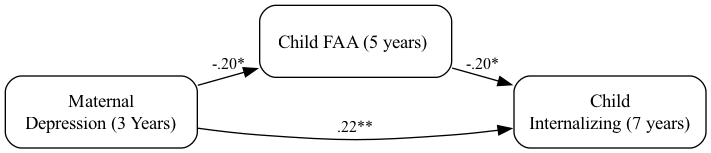

*Note*. Maternal Depression = Revised Beck Depression Inventory (BDI-IA); FAA = EEG Frontal Alpha Asymmetry (log relative); Child Internalizing = Child Behavior Checklist 6-18 years (CBCL).
* *p*<.05, ** *p*<.01

**Supplementary Moderation Analysis**

***Maternal Anxiety Symptoms***

Child FAA at 5 years and maternal anxiety symptoms at 5 years independently contributed to the prediction of child internalizing symptoms at 7 years (*p* = .022 and *p* = .009 respectively). The interaction term (child FAA x maternal anxiety symptoms) was not significant (*p* =.891). Thus, no moderation effect was observed (Supplementary Table 3).

**Supplementary Table 3**

*Regression model for maternal anxiety symptoms (5 years), child FAA (5 years), and child internalizing symptoms (7 years)*

|  | B | B(S) | SE | t | p | LLCI | ULCI |
| --- | --- | --- | --- | --- | --- | --- | --- |
| Maternal Anxiety – 5 Years | 0.32 | .26 | 0.12 | 2.66 | .009** | 0.08 | 0.56 |
| Child FAA – 5 years | -14.79 | -.20 | 6.36 | -2.33 | .022* | -27.26 | -2.32 |
| Maternal Anxiety*Child FAA | 0.11 | .01 | 0.82 | 0.14 | .891 | -1.50 | 1.72 |

*Note.* Maternal Anxiety = Trait Anxiety form of the Spielberger State-Trait Anxiety Inventory (STAI-T); Child FAA = EEG Frontal Alpha Asymmetry (log relative); Child Internalizing = Child Behavior Checklist 6-18 years (CBCL).

***Maternal Depressive Symptoms***

Child FAA at 5 years and maternal depressive symptoms at 5 years independently contributed to the prediction of child internalizing symptoms at 7 years (*p* = .029 and *p* = .002 respectively). The interaction term (child FAA x maternal depressive symptoms) was not significant (*p* =.184). Thus, no moderation effect was observed (Supplementary Table 4).

**Supplementary Table 4**

*Regression model for maternal depressive symptoms (5 years), child FAA (5 years), and child internalizing symptoms (7years)*

|  | B | B(S) | SE | t | p | LLCI | ULCI |
| --- | --- | --- | --- | --- | --- | --- | --- |
| Maternal Depression – 5 Years | 0.64 | .30 | 0.20 | 3.18 | .002** | 0.25 | 1.03 |
| Child FAA – 5 years | -13.79 | -.19 | 6.24 | -2.21 | .029* | -26.02 | -1.55 |
| Maternal Depression*Child FAA | 1.85 | .11 | 1.38 | 1.34 | .184 | -0.86 | 4.55 |

*Note.* Maternal Depression = Revised Beck Depression Inventory (BDI-IA); FAA = EEG Frontal Alpha Asymmetry (log relative); Child Internalizing = Child Behavior Checklist 6-18 years (CBCL).
